# Non-invasive prenatal testing of fetal aneuploidies using a new method based on digital droplet PCR and cell free fetal DNA

**DOI:** 10.1101/2020.12.19.20248553

**Authors:** Wang Haidong, Yang Zhijie, Elena Picchiassi, Federica Tarquini, Giuliana Coata, Wang You, Wang Youxiang, Chen Yu, Gian Carlo Di Renzo

**Author notes:** **Correspondence** Main Corresponding author: Gian Carlo Di Renzo, Department of Medicine and Surgery, Centre of Perinatal and Reproductive Medicine, University of Perugia, Italy., Second Corresponding author: Yu Chen, Department of Obstetrics and Gynecology, The Affiliated Wuxi Maternity and Child Health Care Hospital of Nanjing Medical University, Wuxi, 214000, P.R. China. Wang Haidong and Yang Zhijie made equal contributions in preparing the manuscript therefore are co-first authors for the manuscript.

## Abstract

**Background:** Current next generation sequencing (NGS) and microarray based Non-Invasive Prenatal Tests (NIPT), used for the detection of common fetal trisomies, are still expensive, time consuming and need to be performed in centralized laboratories. To improve NIPT in clinical routine practice as universal prenatal screening, we have developed a digital droplet PCR (ddPCR) based assay called iSAFE NIPT using cell free fetal DNA (cffDNA) for detection of fetal trisomies 13, 18 and 21 in a single reaction with advantage of high diagnostic accuracy and reduced cost.

**Materials and Methods:** We first used artificial DNA samples to evaluate analytical sensitivity and specificity of the iSAFE NIPT. Next, we analysed 269 plasma samples for the clinical validation of iSAFE NIPT. Fifty-eight of these, including five trisomies 21, two trisomies 18 and one trisomy 13 were utilised to establish the assay cut-off values based on ratios between chromosome counts. The remaining 211 plasma samples, including 10 trisomies 21, were analysed to evaluate iSAFE NIPT clinical performance.

**Results:** iSAFE NIPT achieved a 100% analytical sensitivity (95% CI 94.9-100% trisomy 21; 79.4-100% trisomy 18; 73.5-100% trisomy 13) and 100% specificity (95% CI 96.3-100% trisomy 21; 97.6-100% trisomy 18; 97.6-100% trisomy 13). It also achieved a 100% clinical sensitivity and specificity for trisomy 21 detection in the 211 clinical samples (95% CI for sensitivity is 69.1-100%, and 95% CI for specificity is 98.2-100%).

**Conclusions:** The iSAFE NIPT is a highly multiplexed ddPCR based assay for detection of fetal trisomies from maternal blood. Based on clinical validation, the iSAFE NIPT has high diagnostic sensitivity and specificity. It can be decentralized in routine clinical laboratories, is fast, easy to use and economical comparing to current NIPT.

## 1 INTRODUCTION

The discovery of cell free fetal DNA (cffDNA) in maternal blood by Lo et al. in 1997 (1) has opened up new possibilities for non-invasive prenatal diagnosis. Since then, the biological knowledge on cffDNA (2-4) and the rapid development of new molecular biology techniques (5-7) has allowed the development of several non-invasive prenatal tests (NIPT) that are now used in clinical routine to detect the common fetal trisomies 13, 18, 21 (T13, T18, T21) (8-13).

Currently, major NIPT products are all based on next generation sequencing (NGS) or microarray based-test reaching high sensitivity and specificity (14-17). However, these methods usually take place in centralized laboratories, are time-consuming and expensive (18-21). Therefore, to implement NIPT in a large clinical routine as universal screening for fetal T13, T18, T21, there is a need to develop a fast, easy to use and cost-effective method with high diagnostic accuracy and that can be performed in local clinical laboratories.

Several studies demonstrated that the digital PCR (dPCR) is the most powerful method available for an accurate quantification of small amount of DNA (22). In this context, dPCR could have the potential for developing a highly sensitive and reproducible NIPT method. dPCR provides a tool to detect and quantify small amount of DNA by counting amplification signals from single molecules. Some research groups used dPCR to detect fetal T21 for NIPT. These studies (23-24) revealed that the limitation of dPCR for developing NIPT is how to utilize the limited amount of cffDNA to generate enough molecule counts for robust statistical analysis that can differentiate a trisomic fetal sample with a low fetal fraction from an euploid fetal sample. To increase the statistical robustness and simplifies laboratory workflow, the digital droplet PCR (ddPCR) seems to be the best solution to overcome these obstacles due to the possibility of multiplexing, as demonstrated by previous studies (25-27).

Recently, Atila Biosystems has developed an innovative approach, iSAFE NIPT, in order to simultaneously target three fetal chromosomes, 13, 18 and 21 in a single digital reaction using extremely high multiplex ddPCR. This amplify 15 selected target regions that have no known SNP, INDEL or CNV mutations on each of the target chromosomes.

In the present study, we describe the development and the technical validation of the iSAFE NIPT by using a new multiplex ddPCR based NIPT technology. Then, we evaluated the clinical performance of our assay analyzing plasma from pregnant patients.

## 2 MATERIALS AND METHODS

### 2.1 Study design

The study design is shown in the Figure 1. Figure 1A describes the procedures we followed for the development and technical validation of the iSAFE NIPT. We first used in silico calculation in order to optimize the precision parameters needed to differentiate euploid from aneuploid samples. Then, we determined the level of multiplex, number of replicates and the DNA amount required to reach the precision. Technical validations were then performed on artificial euploid (negative) and trisomic (positive) samples. Figure 1B describes the procedures we followed for the clinical validation and test performance of the iSAFE NIPT on 269 plasma samples from pregnant women.

**FIGURE 1.**
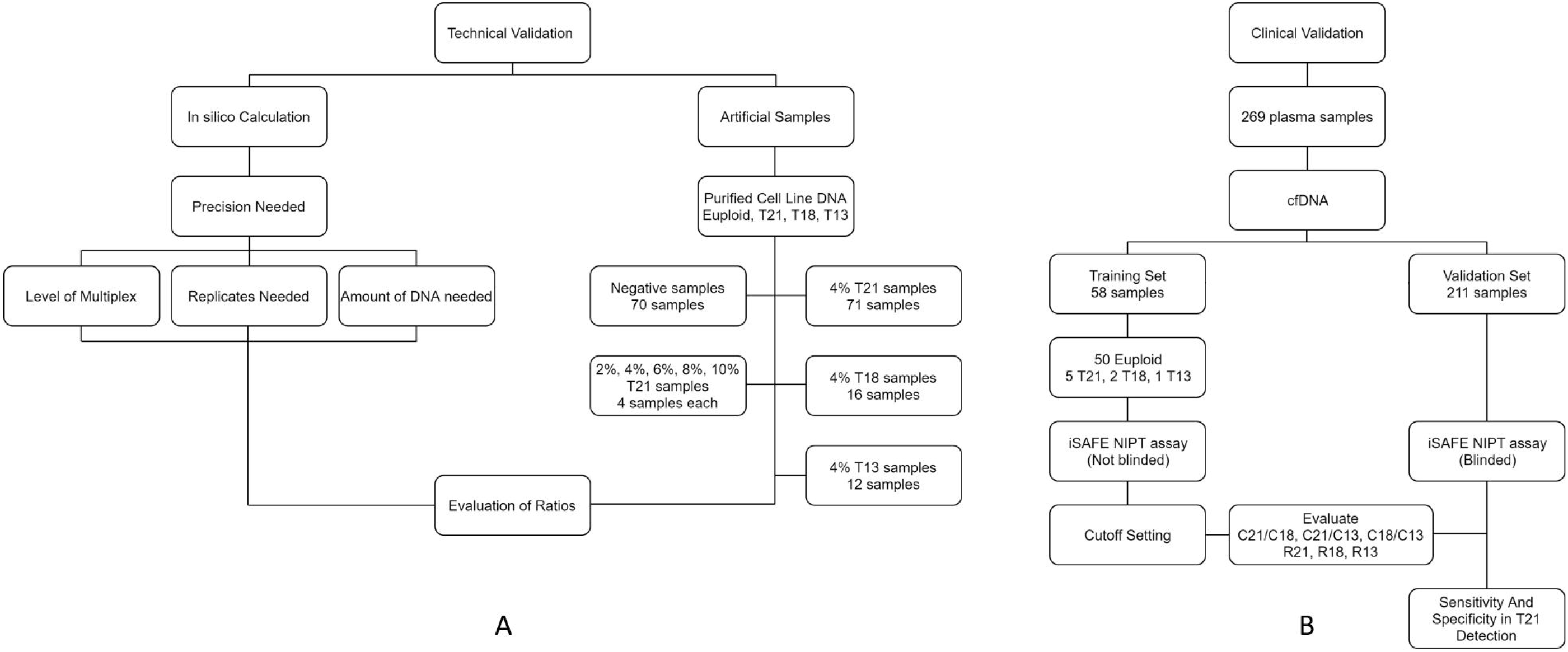
Study design. (A) The procedures for technical validation. (B) The procedures for clinical validation

### 2.2 Technical validation

#### 2.2.1 Preparation of artificial DNA samples

Euploid human genomic DNA was purchased from Millipore (Cat# 69237). DNA of trisomic patients was purchased from Coriell Institute (Cat# NG05121 for T21, Cat# NA02732 for T18, and Cat# 02948 for T13).

Euploid and trisomic genomic DNA samples were digested by CviQI for DNA fragmentation and to improve ddPCR performance. The restriction enzyme digestion was carried out in 40 uL reactions, including 1-4 ug of genomic DNA, 1X NEB buffer 3.1, and 10-40 units of enzyme (CviQI, New England BioLabs, R0639L). The reactions were incubated at room temperature for at least 1 hour. Digested DNA fragments were purified by 2X Beckman Coulter AMPURE XP beads (Cat# A63881) following manufacturer’s manual.

The purified fragmented genomic DNA was quantified by ddPCR using iSAFE NIPT assay. Artificial negative DNA samples were prepared by directly using the fragmented euploid human genomic DNA. Artificial positive DNA samples were prepared by diluting trisomic DNA with euploid DNA sample to obtain the desired percentage of trisomic DNA. The amount of input DNA per sample for ddPCR was 16 ng.

### 2.3 Clinical validation

#### 2.3.1 Plasma DNA samples

A total of 269 pregnant women at 11-27 weeks of gestation were enrolled. Seventy-eight samples were collected from the Obstetrics and Gynecology Outpatient Clinic of the University Hospital of Perugia (Italy) and 191 samples were collected by The Affiliated Huaian No.1 People’s Hospital Nanjing Medical University (China). The study protocol was approved by the Medical Ethics Committee of the Region of Umbria, Italy (CEAS Umbria; approval no. 3352/18) and by the Affiliated Huaian No.1 People’s Hospital Nanjing Medical University, China (Approval no. KY-P-2019-043-01). The experimental procedures adhered to the ethical standards for human experimentation of the 1975 Declaration of Helsinki (revised in 1983).

Peripheral blood samples were collected in EDTA blood collection tubes or Streck tubes (Streck, Cat# CELL-FREE DNA BCT). The blood was centrifuged at 1600g for 10 minutes and the obtained plasma was centrifuged again at 16000g for 10 minutes. Then the plasma was divided into aliquots of 4-6 ml and stored at -80°C until DNA purification.

The confirmation of the ploidy of the fetuses was obtained by CVS, amniocentesis or clinical outcome at birth.

##### Training set samples

During the first non-blinded phase, we analyzed fifty euploid, five T21, two T18 and oneT13. plasma samples to establish the cut-off setting of the iSAFE NIPT. The plasma samples of the training set were selected among recruited pregnant women and the composition of this group was known before ddPCR analysis by laboratory staff.

##### Validation set samples

In the second blinded-phase, we analyzed the remaining 211 plasma samples to assess the iSAFE NIPT diagnostic performance. The composition of this group was unknown before ddPCR by the laboratory staff.

#### 2.3.2 cfDNA purification of plasma samples

cfDNA was extracted from 4-6 mL of maternal plasma samples by QIAamp MinElute ccfDNA Midi Kit (Qiagen 55284) following manufacturer’s instruction. Purified cfDNA was eluted in 40-60 uL TE.

#### 2.3.3 iSAFE NIPT ddPCR assay

iSAFE NIPT reactions were performed in eigth wells for each sample. The 160 uL reaction contained 40 uL of artificial or plasma DNA sample, 16 uL iSAFE NIPT primer and probe mix (Atila Biosystems), 1xBioRad ddPCR™ Supermix for Probes (No dUTP) (BioRad Cat# 1863023), and 4uL iSAFE NIPT buffer I (Atila Biosystems).

The assembled reaction was incubated at room temperature for 5 minutes before droplet generation, and droplets were generated following instruction manual of BioRad Droplet Generator QX200 (BioRad Cat# 186-4002). PCR program was as following: [95°C 10min]X 1, [95°C 15sec and 60°C 3min]X10, [95°C 15sec and 66°C 60sec]X40, and [98 °C 10min]X1. Plates were read in the BioRad QX200 Droplet Reader (BioRad Cat# 186-4001).

ddPCR output was analyzed by BioRad QuantaSoft Analysis Pro (Version 1.0.596). iSAFE NIPT assay was designed to simultaneously quantify chromosome 21, 18, and 13 in a single digital reaction. Two targets (chromosome 21 and 18) were multiplexed in the FAM channel with chromosome 21 being the higher amplitude droplets and chromosome 18 being the lower amplitude droplets, while in HEX channel only one target was counted (chromosome 13).

Two thresholds are drawn in the FAM channel, one between the first and second clusters from bottom to differentiate negative droplets and droplets that are positive for chromosome 21 and/or chromosome 18, and the second one between the second and third clusters to differentiate droplets that are negative for chromosome 21 and droplets that are positive for chromosome 21. One threshold is drawn in the HEX channel between the two clusters to differentiate the positive and negative droplets for chromosome 13.

In ddPCR setting, final data output is the effective number of droplets, i.e. total number of effective droplets, number of positive droplets (with DNA target), and number of negative droplets (without DNA target). The copy number of PCR targets in the ddPCR reaction can be calculated using the equation:

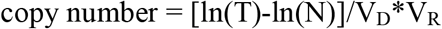

where T is the total number of droplets, N is the number of negative droplets, V_D_ is the mean volume of droplets, and V_R_ is the volume of original ddPCR reaction. Chromosome 18 counts (C18) is calculated by subtraction between the total counts in the FAM channel (by drawing the first threshold) and C21. Chromosome 13 counts (C13) is the count derived in the HEX channel. Chromosome 21 counts (C21) is the count derived in the FAM channel by drawing the second threshold

For artificial samples and plasma samples of training and validation sets, ratios between the three chromosome copy numbers, C21/C18, C21/C13, and C18/C13 were calculated. Furthermore, for plasma samples of training and validation sets three more ratios were calculated R21=[C21/(C18+C13)], R18=[C18/(C21+C13)], and R13=[C13/(C21+C18)].

### 2.4 Statistical analysis

Median and MAD (Median Absolute Deviation) for each ratio were calculated in R: A language and environment for statistical computing, R Core Team (2013).

## 3 RESULTS

### 3.1 Technical validation

#### 3.1.1 iSAFE NIPT assay design

The design of the iSAFE NIPT assay was set up considering the optimal multiplexing, the precision of ddPCR reaction needed to differentiate an euploid from aneuploid sample with 4% fetal fraction, the number of replicates needed for the analysis of each sample and the number of regions to be amplified for each chromosome 21, 18 and 13.

##### The multiplexing

iSAFE NIPT assay was designed to simultaneously detect T21, T18, and T13 in a single digital reaction. Figure 2 shows a typical iSAFE NIPT assay ddPCR output generated by performing the iSAFE NIPT assay on 2 ng fragmented human genomic DNA. The clear separation between the clusters is essential for accurate estimation of the copy number of the chromosomes.

**FIGURE 2.**
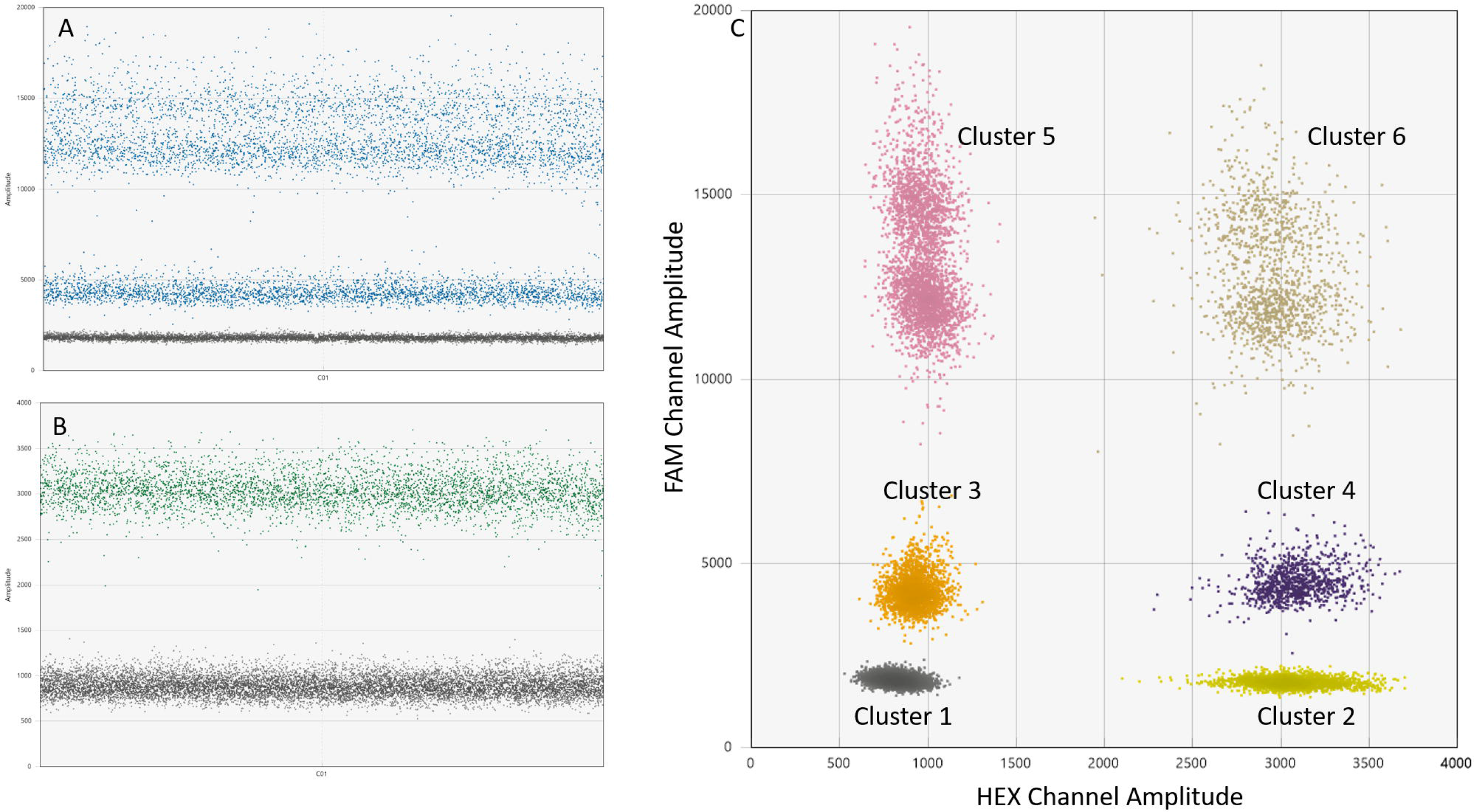
1D and 2D figures of iSAFE NIPT assay ddPCR output. (A) 1D figure of the FAM channel. Three clusters are observed in the FAM channel. From bottom to top, the three clusters are negative droplets, chromosome 18 positive droplets, chromosome 21 or chromosome 21/ chromosome 18 positive droplets. (B) 1D figure of the HEX channel. Two clusters are observed in the HEX channel. From bottom to top, the two clusters are negative droplets, chromosome 13 positive droplets. (C) 2D figure of the ddPCR output showing 6 clusters. Cluster 1 (gray) is triple-negative droplets; cluster 2 (yellow) is chromosome 13 positive droplets; cluster 3 (orange) is chromosome 18 positive droplets; cluster 4 (purple) is C18/C13 double positive droplets; cluster 5 (pink) is chromosome 21 positive droplets and chromosome 21/ chromosome 18 double positive droplets; and cluster 6 (brown) is chromosome 21/ chromosome 13 double positive droplets and chromosome 21/ chromosome 18/ chromosome 13 triple positive droplets

##### The precision of ddPCR reaction needed to differentiate a euploid from aneuploid sample

We used a low fetal fraction at 4% for precision evaluation. In order to differentiate a euploid from an aneuploid sample with 4% fetal fraction, a 99% confidence would require the precision (here we use CV%=Standard deviation/Mean*100% for the precision) to be at 0.86%, the required CV% is calculated by FF%/2/Z_1-confidence_. The precision of a ddPCR reaction is determined by copy number of PCR targets and total number of droplets. The following equations (28) were used in the calculation, where 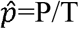 (P is the number of positive droplets and T is the total number of droplets), *Z*_*α*_ is the Z value for the *α* for a corresponding confidence level.

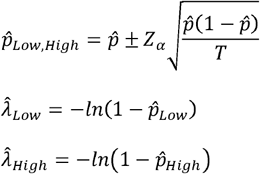

The precision or the CV% is then calculated by 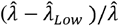 and 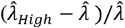 using *Z*_*α*_=1. As shown in the equations, the precisions are slightly different on the two ends and for convenience in description we use the average value to express the precision. The best precision (the lowest CV%) is achieved when 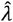 is at ∼1.6 (29). For any single ddPCR well reaction, total number of droplets can vary from 10000 to 20000. For example, if we use the low end value 10000 in the calculation, the best precision is at ∼1.24% for one well reaction, ∼0.88% for two well reaction, ∼0.62% for four well reaction, ∼0.51% for six well reaction, and ∼0.44% for eight well reaction. one or two well reactions would not be ideal in the situation. The number of PCR targets required would be ∼16900 for four well reaction, ∼15500 for six well reaction, and ∼14900 for eight well reactions to reach a precision at 0.86%. 10000 droplets would mean that dead reaction volume is over 50%, the number of required PCR targets should be multiplied by two at least.

##### The number of well reactions needed for analysis of each sample

Another factor to be considered is the number of well reactions needed for the analysis of each sample. 16900 targets in four well reaction results in a CPD at 1.27 (16900*3/40000) for three chromosomes. Similarly, 15500 targets in six well reaction results in a CPD at 0.78, and the number is below 0.56 for 14900 targets in eight or more well reactions. At CPD 0.56, majority of positive droplets (more than 99%) contain 1-4 targets, a few positive droplets can contain five targets and very rarely a droplet can contain six targets simultaneously. When the CPD increases to 0.78 or 1.28, 7 targets or eight targets in a single droplet start to show up. In the current study, eight well reactions are preferred over four or six wells for the iSAFE NIPT assay because: 1) the competition between different types of target is lower when using eight wells; 2) it has better assay precision due to more droplets are generated; 3) further increasing the number of reactions would increase cost, make experimental procedure less convenient and reduce assay throughput.

##### The number of regions to be amplified for each target chromosome

Based on the above information, we used 14900 targets in eight well reactions, or 29800 targets if we consider the ddPCR dead volume, to assess the number of regions to be amplified for each of the chromosome 21, 18 and 13. Average whole 20 ng cfDNA can be purified from a 4 mL typical maternal blood plasma sample or 10mL of maternal blood sample. Knowing that one ng of cfDNA contain about 300 target copies, if we use a low end 10 ng in the calculation, 29800 copy targets requires at least ∼29800/(10*300) ∼ 10 targets per chromosome. Therefore, we selected 15 regions as PCR targets for each of the three chromosomes.

#### 3.1.2 iSAFE NIPT assay validation by artificial DNA samples

ddPCR results obtained from serial dilutions of artificial DNA T21 samples are shown in Table 1. Three ratios (C21/C18, C21/C13, and C18/C13) are reported for each dilution in four replicates. Student t-Test was performed on the three ratios between the positive and negative samples and p values are shown in the table. For all artificial T21 samples, both ratios C21/C18 and C21/13 showed significant p values (<=0.05) while ratio C18/C13 showed non-significant p values. When we set 1.01 as cut-off value, both ratios C21/C18 and C21/C13 in all artificial T21 samples were higher than the cut-off. Although ratio C21/C18 of sample 3 in the artificial negative samples is larger than 1.01, since ratio C21/C13 of the same sample is smaller than 1.01, the sample 3 is still categorized as a negative sample.

**TABLE 1.**
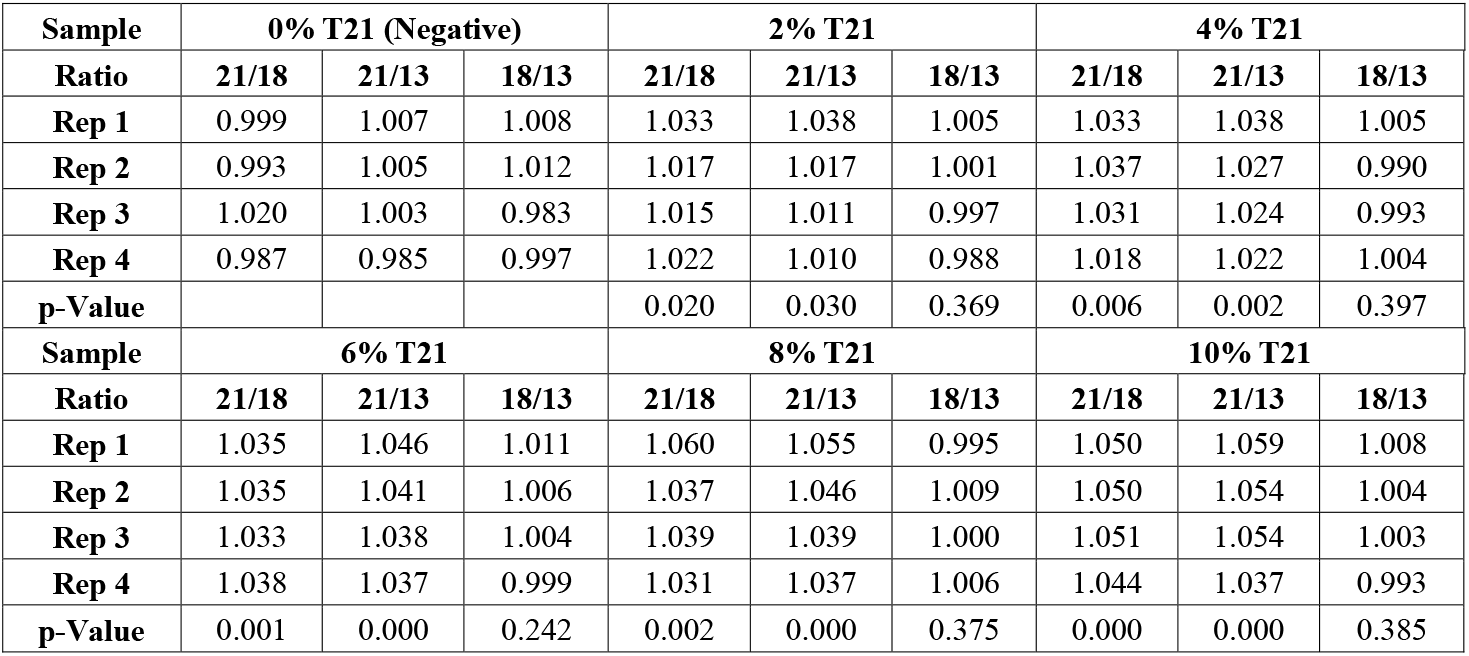
Chromosome ratio values in the artificial euploid and aneuploid samples. The aneuploid samples include serial dilutions of trisomic patient DNA in euploid DNA. Each sample is done in 4 replicates

| **Sample** | **0% T21 (Negative)** |  |  | **2% T21** |  |  | **4% T21** |  |  |
| --- | --- | --- | --- | --- | --- | --- | --- | --- | --- |
| **Ratio** | **21/18** | **21/13** | **18/13** | **21/18** | **21/13** | **18/13** | **21/18** | **21/13** | **18/13** |
| **Rep 1** | 0.999 | 1.007 | 1.008 | 1.033 | 1.038 | 1.005 | 1.033 | 1.038 | 1.005 |
| **Rep 2** | 0.993 | 1.005 | 1.012 | 1.017 | 1.017 | 1.001 | 1.037 | 1.027 | 0.990 |
| **Rep 3** | 1.020 | 1.003 | 0.983 | 1.015 | 1.011 | 0.997 | 1.031 | 1.024 | 0.993 |
| **Rep 4** | 0.987 | 0.985 | 0.997 | 1.022 | 1.010 | 0.988 | 1.018 | 1.022 | 1.004 |
| **p-Value** |  |  |  | 0.020 | 0.030 | 0.369 | 0.006 | 0.002 | 0.397 |
| **Sample** | **6% T21** |  |  | **8% T21** |  |  | **10% T21** |  |  |
| **Ratio** | **21/18** | **21/13** | **18/13** | **21/18** | **21/13** | **18/13** | **21/18** | **21/13** | **18/13** |
| **Rep 1** | 1.035 | 1.046 | 1.011 | 1.060 | 1.055 | 0.995 | 1.050 | 1.059 | 1.008 |
| **Rep 2** | 1.035 | 1.041 | 1.006 | 1.037 | 1.046 | 1.009 | 1.050 | 1.054 | 1.004 |
| **Rep 3** | 1.033 | 1.038 | 1.004 | 1.039 | 1.039 | 1.000 | 1.051 | 1.054 | 1.003 |
| **Rep 4** | 1.038 | 1.037 | 0.999 | 1.031 | 1.037 | 1.006 | 1.044 | 1.037 | 0.993 |
| **p-Value** | 0.001 | 0.000 | 0.242 | 0.002 | 0.000 | 0.375 | 0.000 | 0.000 | 0.385 |

The data on 70 artificial negative samples and 71 artificial positive T21 samples with T21 DNA at 4% are summarized in Table 2. We could again apply the cutoff value at 1.01 to differentiate negative and positive samples. We performed similar experiments on 16 artificial positive T18 samples with T18 DNA at 4% and 12 artificial positive T13 samples with T13 DNA at 4% (Table 2). The cutoff values to differentiate negative and positive T18 samples were 0.985 for C21/C18 and 1.021 for C18/C13. The cutoff values to differentiate negative and positive T13 samples were 0.982 for C21/C13 and 0.985 for C18/13.

**TABLE 2.**
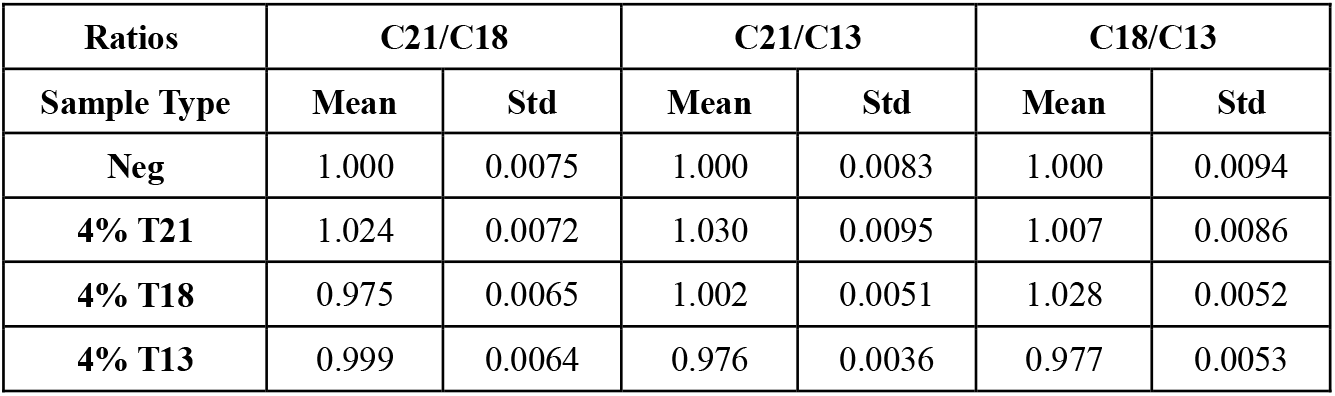
Means and standard deviations of chromosome ratio in the artificial euploid and aneuploid samples for variability evaluation

| **Ratios** | **C21/C18** |  | **C21/C13** |  | **C18/C13** |  |
| --- | --- | --- | --- | --- | --- | --- |
| **Sample Type** | **Mean** | **Std** | **Mean** | **Std** | **Mean** | **Std** |
| **Neg** | 1.000 | 0.0075 | 1.000 | 0.0083 | 1.000 | 0.0094 |
| **4% T21** | 1.024 | 0.0072 | 1.030 | 0.0095 | 1.007 | 0.0086 |
| **4% T18** | 0.975 | 0.0065 | 1.002 | 0.0051 | 1.028 | 0.0052 |
| **4% T13** | 0.999 | 0.0064 | 0.976 | 0.0036 | 0.977 | 0.0053 |

### 3.2 Clinical validation

#### 3.2.1 iSAFE NIPT assay on plasma samples of training set

The training set contained 50 euploid samples, five T21 samples, two T18 samples, and one T13 sample. These samples were used to set the cut-off values to distinguish euploid from aneuploid fetuses. The three pairwise ratios obtained between the three chromosomes, C21/C18, C21/C13, and C18/C13 for each sample from the training set are shown in the Table 3A.

**TABLE 3.** Range, median, and MAD values for the chromosome ratios in clinical training sample set and validation sample set. (A) Ratio C21/C18, Ratio C21/C13, and Ratio C18/C13 in the training sample set; (B) Ratio R21, R18, and R13 in the training sample set; (C) Ratio C21/C18, Ratio C21/C13, and Ratio C18/C13 in the validation sample set; (D) Ratio R21, R18, and R13 in the validation sample set

| **A** | **C21/C18** |  |  | **C21/C13** |  |  | **C18/C13** |  |  |
| --- | --- | --- | --- | --- | --- | --- | --- | --- | --- |
|  | **Range** | **Median** | **MAD** | **Range** | **Median** | **MAD** | **Range** | **Median** | **MAD** |
| **Neg** | 0.965-1.038 | 1.003 | 0.022 | 0.964-1.046 | 1.003 | 0.024 | 0.954-1.052 | 0.999 | 0.015 |
| **T21** | 1.047-1.086 | 1.065 | 0.006 | 1.047-1.133 | 1.067 | 0.030 | 0.994-1.061 | 1.004 | 0.009 |
| **T18** | 0.930-0.944 | 0.937 | 0.010 | 1.017-1.019 | 1.018 | 0.002 | 1.081-1.095 | 1.088 | 0.010 |
| **T13** | 1.052 |  |  | 0.952 |  |  | 0.906 |  |  |

| **B** | **R21** |  |  | **R18** |  |  | **R13** |  |  |
| --- | --- | --- | --- | --- | --- | --- | --- | --- | --- |
|  | **Range** | **Median** | **MAD** | **Range** | **Median** | **MAD** | **Range** | **Median** | **MAD** |
| **Neg** | 0.969-1.033 | 1.002 | 0.019 | 0.960-1.040 | 1.000 | 0.015 | 0.961-1.030 | 0.997 | 0.021 |
| **T21** | 1.048-1.102 | 1.066 | 0.027 | 0.965-0.996 | 0.972 | 0.009 | 0.911-0.977 | 0.966 | 0.017 |
| **T18** | 0.974-0.982 | 0.978 | 0.006 | 1.071-1.086 | 1.078 | 0.011 | 0.948-0.953 | 0.951 | 0.004 |
| **T13** | 0.999 |  |  | 0.928 |  |  | 1.077 |  |  |

| **C** | **C21/C18** |  |  | **C21/C13** |  |  | **C18/C13** |  |  |
| --- | --- | --- | --- | --- | --- | --- | --- | --- | --- |
|  | **Range** | **Median** | **MAD** | **Range** | **Median** | **MAD** | **Range** | **Median** | **MAD** |
| **Neg** | 0.948-1.062 | 1.000 | 0.018 | 0.947-1.045 | 1.000 | 0.017 | 0.947-1.051 | 1.000 | 0.018 |
| **T21** | 1.027-1.128 | 1.097 | 0.037 | 1.048-1.127 | 1.073 | 0.029 | 0.960-1.021 | 1.000 | 0.023 |

| **D** | **R21** |  |  | **R18** |  |  | **R13** |  |  |
| --- | --- | --- | --- | --- | --- | --- | --- | --- | --- |
|  | **Range** | **Median** | **MAD** | **Range** | **Median** | **MAD** | **Range** | **Median** | **MAD** |
| **Neg** | 0.950-1.037 | 1.000 | 0.016 | 0.945-1.039 | 1.000 | 0.015 | 0.951-1.043 | 1.000 | 0.015 |
| **T21** | 1.038-1.123 | 1.082 | 0.038 | 0.924-0.998 | 0.954 | 0.029 | 0.935-0.990 | 0.966 | 0.016 |

A threshold could be drawn for C21/C18 between 1.038 and 1.047 to differentiate euploid from T21 samples and another threshold could be drawn between 0.965 and 0.944 to differentiate euploid from T18 samples. Similarly, for C21/C13, a threshold at 1.046 can differentiate euploid from T21 samples, a threshold between 0.964 and 0.952 can differentiate euploid and T13 samples. For C18/C13, a threshold between 1.052 and 1.081 can differentiate euploid and T18 samples, a threshold between 0.954 and 0.906 can differentiate euploid and T13 samples. To improve the performance of the test, we explored if two chromosomes combined can be used as reference to evaluate the copy number of the third chromosome. Therefore, other three ratios R21=[C21/(C18+C13)], R18=[C18/(C21+C13)] and R13=[C13/(C21+C18)] were calculated using the same results from the training set. The ratio values were normalized to 1 for convenience and are shown in Table 3B.

A threshold could be drawn for R21 between 1.033 and 1.048 to differentiate euploid from T21 samples. A threshold could be drawn for R18 between 1.040 and 1.071 to differentiate euploid from T18 samples. Finally, a threshold could be drawn for R13 between 1.030 and 1.077 to differentiate euploid from the T13 sample.

#### 3.2.2 iSAFE NIPT assay on plasma samples of validation set

The validation study was performed only for T21 since we did not have any T18 and T13 samples in the validation sample set.

Ranges, median, and MAD values for C21/C18, C21/C13 and C18/C13 are summarized in the Table 3C, and the values for R21, R18, and R13 are summarized in the Table 3D. Boxplots, beeswarm plots (Figure 3) and three receiver operating characteristics (ROC) curves (Figure 4) are plotted for T21 detection by C21/C18, C21/C13 and R21, respectively. C21/C13 and R21 showed nearly perfect ROC curves and C21/C18 curve also shows the shape of a particularly good screening test.

**FIGURE 3.**
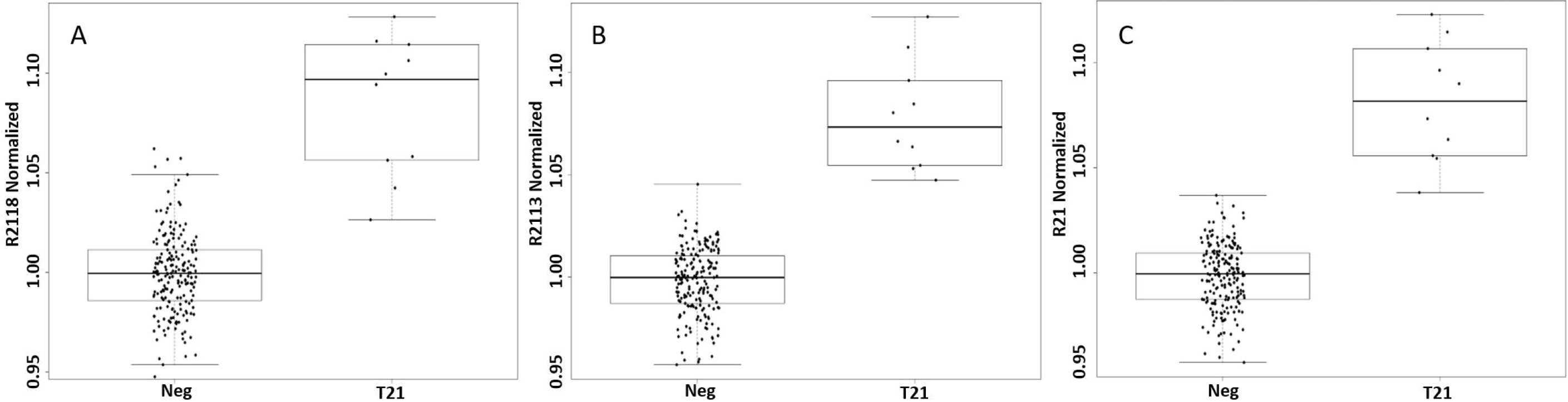
iSAFE NIPT data of clinical samples in the format of boxplot and beeswarm plot. (A) The distribution of ratio C21/C18 in negative samples and T21 samples. (B) The distribution of ratio C21/C13 in negative samples and T21 samples. (C) The distribution of ratio R21 in negative samples and T21 samples. C21 is the chromosome 21 counts, C18 is the chromosome 18 counts, and C13 is the chromosome 13 counts. R21, R18, R13 are calculated as R21=[C21/(C18+C13)], R18=[C18/(C21+C13)], and R13=[C13/(C21+C18)]

**FIGURE 4.**
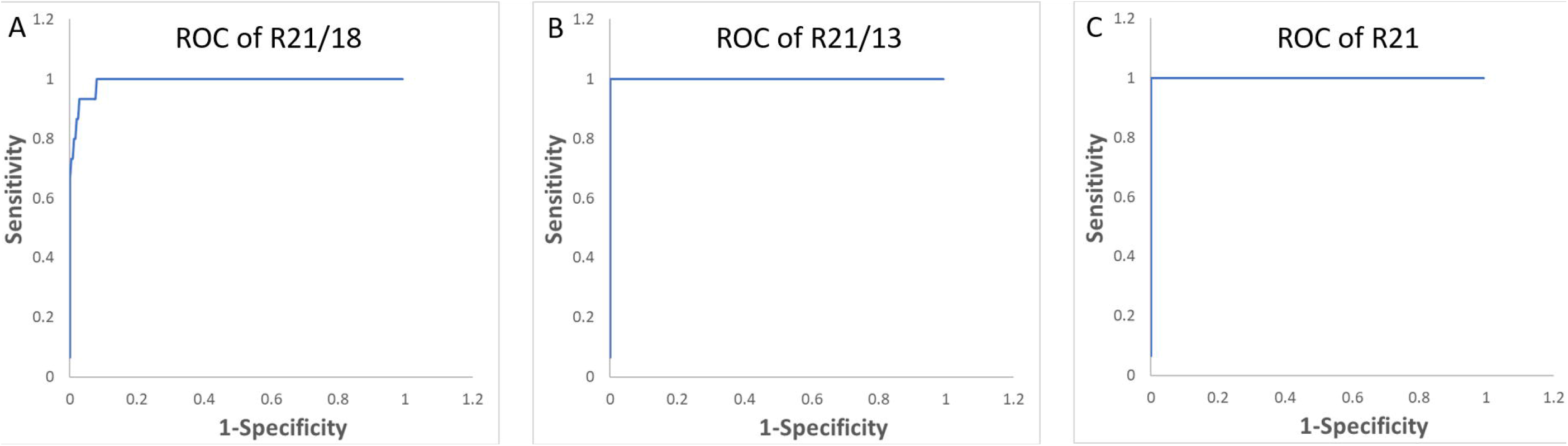
The three ROC curves for trisomy 21 detection, using (A) C21/C18 as a predictor; C21/C13 as a predictor; (C) R21 as a predictor

Based on the ROC curves, using C21/C18, a threshold at 1.038 correctly classifies 9 T21 samples out of a total of 10 T21 samples (sensitivity at 90%), and correctly classifies 193 euploid samples out of a total of 201 euploid samples (specificity at 96.0%). A threshold at 1.047 correctly classifies 8 T21 samples (sensitivity at 80%), and correctly classifies 196 euploid samples (specificity at 97.5%). A threshold at 1.046 correctly classified all euploid samples and all T21 samples (sensitivity and specificity at 100%).

Using R21, a threshold at 1.033 correctly classifies all T21 samples (sensitivity at 100%), and mis-classifies one euploid sample as T21 sample (specificity at 99.5%). A threshold at 1.048 mis-classifies one T21 sample as euploid sample (sensitivity at 90%) and correctly classifies all euploid samples (specificity at 100%). However, a threshold at 1.037 can bring both sensitivity and specificity to 100%.

## 4 DISCUSSION

In this study, we described and validated a new method of NIPT for the detection of fetal trisomies based on high multiplexed ddPCR and cffDNA from maternal plasma.

The current effort is to develop a ddPCR based NIPT assay that can be comparable, in term of performance, to the NIPT assays based on NGS or microarray platforms. Similar studies have used dPCR platform for the detection of fetal chromosomal aneuploidies and have proven the feasibility and reliability of dPCR based assays. Khattabi et al. used chromosome 1 as the reference in T21 detection and has stated the possibility that both T21 and T18 can be detected when PCR targets are selected on the two chromosomes (25). This has been then realized by Tan et al. using chromosome 21 and chromosome 18 as references to each other (27).

Our study further added chromosome 13 probes and is the first test that simultaneously target three chromosomal aneuploidies in one digital PCR assay with increased multiplicity level. The chromosome 21 and chromosome 18 positive droplets were differentiated in the FAM channel by amplitude, where chromosome 21 positive droplets had higher amplitude and chromosome 18 positive droplets had lower amplitude. The chromosome 13 positive droplets were detected in the HEX channel.

In the clinical validation, three indicators, C21/C13, C21/C18, and R21, were used to draw cut- off values in order to differentiate T21 samples and euploid samples. We demonstrated that using either the ratio C21/C13 or the ratio R21 as indicator, our assay could differentiate T21 and euploid samples and reached both 100% sensitivity and 100% specificity in the current sample set. Using the ratio C21/C18, the sensitivity was 90% and the specificity was 96% for T21 detection. We speculated that Chromosome 18 positive droplets were located in the FAM channel at lower amplitude and the counts of Chromosome 18 was a derived value from the subtraction of direct measurement of C21+C18 and direct measurement of C21, which results in increased variability.

As new generations of digital PCR devices will be able to accommodate more fluorescence channels, additional chromosomes can be targeted for more aneuploidy detection, as well as that assay precision can be further improved by detecting single chromosome per channel.

In summary, C21/C13 or R21 are reliable indicators for T21 detection. More samples can be tested in order to determine if the ratio C21/C18 is a reliable indicator, and if a test based on both C21/C18 and C21/C13 would be a more powerful test than C21/C13 alone. The clinical samples for validation in this study do not have T18 or T13 samples due to the rareness of these aneuploidies, the clinical performance of these two aneuploidies need to be confirmed in a larger cohort.

The fetal fraction of cfDNA is an important quality control factor in the NIPT assay, as the amount of aneuploid chromosomal material increases linearly as fetal fraction increases. Atila Biosystems has developed a general fetal fraction assay for both male and female fetus, and a gender determination assay which reports gender and fetal fraction for male fetus. Both assays can be performed under the same condition as the iSAFE NIPT assay, they can be combined in the same ddPCR run. The development and validation of the fetal fraction assays will be reported in a separate study.

The cost of current NGS and microarray based NIPT tests are still high (18-21). In many countries, NIPT test is not covered by insurance and entire cost is paid by customers out-of-pocket. Due to large manpower and equipment investments required by setting up and running a NIPT specialized laboratory, complicated sample process and time-consuming data analysis, these tests are only provided by centralized laboratories. On the contrary, iSAFE NIPT provides significantly simpler workflow and therefore can be installed and performed in decentralized local clinical laboratories. A single professional can manage the whole workflow and obtain the results within 2.5 hours for a single sample, as the iSAFE NIPT not required to be performed in batch, data analysis is straight forward and readily finished on regular laboratory equipped computers. The key advantage is that the iSAFE NIPT is a digital PCR based test that directly takes cfDNA as sample input and does not require any sample processing, such as the NGS library preparation. The throughput of the iSAFE NIPT assay is 12 samples per 96 well PCR plate. Using the BioRad ddPCR systems (www.bio-rad.com) as an example, with one PCR thermocycler and one droplet reader, 36-48 samples can be processed daily. The hands-on time can be further reduced if an automated droplet generator is available.

In conclusion, iSAFE NIPT has a great potential as universal prenatal screening of the common fetal trisomies and it could be implemented as a clinical routine practice and extended to all the pregnant women in any part of the world.

## Data Availability

All data is available.

## ACKNOWLEDGMENTS

This study was supported by the University of Perugia (Italy) Fondo Ricerca di Base 2018.

## REFERENCES

1. Lo YM, Corbetta N, Chamberlain PF, Rai V, Sargent IL, Redman CW, Wainscoat JS. Presence of fetal DNA in maternal plasma and serum. Lancet 1997;350(9076):485–487.

2. Lo YM, Tein MS, Lau TK et al. Quantitative analysis of fetal DNA in maternal plasma and serum: implications for noninvasive prenatal diagnosis. Am J Hum Genet. 1998;62(4):768–775.

3. Bischoff FZ, Lewis DE, Simpson JL. Cell-free fetal DNA in maternal blood: kinetics, source and structure. Hum Reprod Update 2005;11:59–67.

4. Chan KC, Zhang J, Hui AB et al. Size distributions in maternal and fetal DNA in maternal plasma. Clin Chem. 2004; 50(1):88–92.

5. Fan HC, Blumenfeld YJ, Chitkara U et al. Noninvasive diagnosis of fetal aneuploidy by shotgun sequencing DNA from maternal blood. Proc. Natl. Acad. Sci USA 2008;105:16266–16271.

6. Chiu RWK, Chan KCA, Gao Y et al. Noninvasive prenatal diagnosis of fetal chromosomal aneuploidy by massively parallel genomic sequencing of DNA in maternal plasma. Proc. Natl. Acad. Sci USA 2008;105:20458–20463.

7. Sparks AB, Struble CA, Wang ET et al. Noninvasive prenatal detection and selective analysis of cell-free DNA obtained from maternal blood: evaluation for trisomy 21 and trisomy 18. Am J Obstet Gynaecol 2012;206:319e1–9.

8. Chiu RW, Akolekar R, Zheng YW et al. Non invasive prenatal assessment of trisomy 21 by multiplexed maternal plasmaDNA sequencing: large scale validity study. BMJ 2011;342:c7401.

9. Norton ME, Jacobssonc B, Swamy GK et al. Cell-free DNA analysis for non invasive examination of trisomy. N. Engl. J. Med. 2015;372(17:1589–1587.

10. Bianchi DW, Parker RL, Wentworth J et al. CARE Study Group. DNA sequencing versus standard prenatal aneuploidy screening. N Engl J Med 2014;370(9):799–808.

11. McCullough RM, Almasri EA, Guan X, et al. Non-invasive prenatal chromosomal aneuploidy testing--clinical experience: 100,000 clinical samples. PLoS One. 2014;9(10):e109173.

12. Taneja PA, Snyder HL, de Feo E, et al. Noninvasive prenatal testing in the general obstetric population: clinical performance and counseling considerations in over 85,000 cases. Prenat Diagn. 2016;36(3):237–243.

13. Samura O. Update on non invasive prenatal testing: a review based on current worldwide research. J Obstet Gynaecol Res 2020;8:1146–1254.

14. Zimmermann B, Hill M, Gemelos G, et al. Non-invasive prenatal aneuploidy testing at chromosomes 13, 18, 21, X, and Y, using targeted sequencing of polymorphic loci. Prenat Diagn. 2012;32(13):1233–1241.

15. Agarwal A, Sayres LC, Cho MK, Cook-Deegan R, Chandrasekharan S. Commercial Landscape of noninvasive prenatal testing in the United States. Prenat Diagn. 2013;33(6):521–53.

16. Juneau K, Bogard PE, Huang S et al. Microarray-based cell-free DNA analysis improves non invasive prenatal testing Fetal Diagn. Ther. 2014;36:282–286.

17. Stokowski R, Wang E, White K et al. Clinical performance of non-invasive prenatal testing (NIPT) using targeted cell-free fetal DNA analysis in maternal plasma with microarray or next generation (NGS) is consistent across multiple controlled clinical studies: clinical performance of targeted cfDNA NIPT. Prenat. Diagn. 2015:35:1243–1246.

18. Cuckle H, Benn P, Pergament E. Maternal CFDNA screening for Down syndrome-a cost sensitivity analysis. Prenat Diagn 2013; 33:636–642.

19. Evans M, Sonek JD, Hallahan TW et al. Cell-free fetal DNA screening in the USA: a cost analysis screening strategies. Ultrasound Obstet Gynecol 2015; 45(1):74–83.

20. Zhang W, Mohammadi T, Sou J et al. Cost-effectiveness of prenatal screening and diagnostic strategies for Down syndrome: a microsimulation modeling analysis. PloS One 2019; 14(12);e0225281.

21. Xu Y, Wei Y, Li N et al. Cost-effectiveness analysis of non-invasive prenatal testing for Down syndrome in China. Int J Technol Assess Health Care 2019; 35(3):237–242.

22. Nectoux J. Current, emerging, and future applications of digital PCR in non-invasive prenatal diagnosis. Mol Diagn Ther 2018; 22:139–148.

23. Lo YMD, Lun FMF, Chan KCA, et al. Digital PCR for the molecular detection of fetal chromosomal aneuploidy. Proc Natl Acad Sci U S A. 2007;104(32):13116–13121.

24. Fan HC, Blumenfeld YJ, El-Sayed YY, Chueh J, Quake SR. Microfluidic digital PCR enables rapid prenatal diagnosis of fetal aneuploidy. Am J Obstet Gynecol. 2009;200(5):543.e1-7.

25. El Khattabi LA, Rouillac-Le Sciellour C, Le Tessier D, et al. Could Digital PCR Be an alternative as a Non-Invasive Prenatal Test for Trisomy 21: A Proof of Concept Study. PLoS One 2016;11(5):e0155009. doi:10.1371/journal.pone.0155009.eCollection 2016

26. Lee SY, Kim SJ, Han S-H, et al. A new approach of digital PCR system for non-invasive prenatal screening of trisomy 21. Clin Chim Acta Int J Clin Chem. 2018;476:75–80.

27. Tan C, Chen X, Wang F, et al. A multiplex droplet digital PCR assay for non-invasive prenatal testing of fetal aneuploidies. Analyst. 2019;144(7):2239–2247.

28. Dube S, Qin J, Ramakrishnan R. Mathematical analysis of copy number variation in a DNA sample using digital PCR on a nanofluidic device. PloS One. 2008;3(8):e2876.

29. Majumdar N, Wessel T, Marks J. Digital PCR Modeling for Maximal Sensitivity, Dynamic Range and Measurement Precision. PloS One. 2015;10(3):e0118833.

